# ^1^H-NMR metabolomics-based surrogates to impute common clinical risk factors and endpoints

**DOI:** 10.1101/2021.07.19.21258470

**Authors:** D. Bizzarri, M.J.T. Reinders, M. Beekman, P.E. Slagboom, BBMRI-NL, E.B. van den Akker

## Abstract

Missing or incomplete phenotypic information can severely deteriorate the statistical power in epidemiological studies. High-throughput quantification of small-molecules in bio-samples, i.e. ‘metabolomics’, is steadily gaining popularity, as it is highly informative for various phenotypical characteristics. Here we aim to leverage metabolomics to impute missing data in clinical variables routinely assessed in large epidemiological and clinical studies. To this end, we have employed ∼25,000 ^1^H-NMR metabolomics samples from 28 Dutch cohorts collected within the BBMRI-NL consortium, to create 19 metabolomics-based predictors for clinical variables, including diabetes status (AUC_5-Fold CV_ = 0.94) and lipid medication usage (AUC_5-Fold CV_ = 0.90). Subsequent application in independent cohorts confirmed that our metabolomics-based predictors can indeed be used to impute a wide array of missing clinical variables from a single metabolomics data resource. In addition, application highlighted the potential use of our predictors to explore the effects of totally unobserved confounders in omics association studies. Finally, we show that our predictors can be used to explore risk factor profiles contributing to mortality in older participants. To conclude, we provide ^1^H-NMR metabolomics-based models to impute clinical variables routinely assessed in epidemiological studies and illustrate their merit in scenarios when phenotypic variables are partially incomplete or totally unobserved.

## INTRODUCTION

A major goal in biomedical research is to find faithful biomarkers of health, defined as accurate and reproducible assays that provide objective indications on the health of an individual and his/her risk of developing a disease over predefined time trajectory [1]. Over the years, many types of putative biomarkers have been proposed, ranging from environmental factors to biochemical assays, that may aid the diagnosis and prognostication of disease, including cardiovascular disease, cancer and immunological disorders. Many of these clinical variables, however, are costly or cumbersome to obtain, especially for more critical and frail participants, such as older individuals [2], [3]. Consequently, missing data frequently occurs in large epidemiological or clinical studies, potentially leading to a significant loss of statistical power, thus impeding biomarker research in studies of older individuals [4].

Missing phenotypic data can be handled in various ways. Often, analyses are either restricted to individuals or variables with complete data, which both may introduce potential biases [5]. Alternatively, missing data can be imputed using complete phenotypic variables [4], [6]–[8], yet these approaches work only satisfactory if the complete phenotypic variables are informative for the ones with missing observations. A third solution basically extents the second approach by leveraging informative omics data to impute missing phenotypic data. Particularly useful in this context are metabolite quantifications in minimally invasive biomaterials, such as urine, saliva or blood plasma, obtained with proton Nuclear Magnetic Resonance (^1^H-NMR) assays [9]. Although this technique only captures a modest number of analytes, ^1^H-NMR metabolomics data is frequently acquired in large-scale epidemiological studies, as it is a cost-efficient and reproducible data resource. The underlying motivation is that metabolite concentrations in blood seem to be direct readouts of various biological processes, incorporating cues of the environment as well as the host’s genetic background, and hence may be regarded as intermediate phenotypes. Indeed, metabolomics has been shown to capture information on the effect of drug treatments [10], disease status [11]–[14], functional and cognitive decline [15], and aging [16], [17]. In addition, several studies used the blood metabolome to predict single anthropometric measures, i.e. BMI [18], or other physiological characteristics, i.e. sex [19] or age [16]. However, it remains unclear whether the blood metabolome captured by ^1^H-NMR could represent phenotypic information over a wider set of conventional clinical variables.

We hypothesize that a single set of blood metabolic markers combined in multiple algorithms may represent a range of conventional clinical variables. As a proof of concept, we generated metabolic surrogates for 20 variables of general clinical and epidemiological interest available in at least 6 of the cohorts collaborating in BBMRI-NL. Here we will designate these as conventional clinical variables and they comprehend physiological measures (sex, age, blood pressure, etc.), environmental exposures (current smoking, etc.), body composition measures (BMI, etc.), inflammatory factors (hsCRP), medication usage (lipids medication, etc.), blood composition (white cell counts, etc.) lipids metabolism (LDL-cholesterol, etc.) and cardiometabolic clinical endpoints (diabetes and metabolic syndrome). Acquiring data for all these variables is costly and requires sufficient biomaterial, meaning that not every study has collected the same set of data. We further explored these methods to establish metabolic surrogate values in the Leiden Longevity Study, which we used to showcase possible applications in epidemiological research. We showed the validity of the surrogates in an external cohort comparing them to the original values, we examined their association to further clinically valuable cardiometabolic health markers, and explored whether the metabolic surrogates associate, separately or combined, to all-cause mortality.

## RESULTS

### 1H-NMR metabolomics can be used to successfully predict 19 out of 20 clinical variables routinely measured in epidemiological and clinical studies

Missing or incomplete phenotypic information can severely deteriorate the statistical power in epidemiological studies. Here we evaluate the ability of Nuclear Magnetic Resonance (^1^H-NMR) metabolomics (Nightingale Health^©^, Helsinki, Finland) to reconstruct conventional clinical variables. For this purpose, we trained and evaluated prediction models (**Figure 1A**) for 20 conventional clinical variables (**Figure 1B**) using data of ∼31,000 individuals collected within the Dutch Biobanking and BioMolecular resources and Research Infrastructure (BBMRI-NL: https://www.bbmri.nl/). Out of 220 metabolomic variables measured on the platform, we employed 56 metabolic markers, selected to be the most uncorrelated [21], [22] and most successfully measured in the BBMRI studies (**Methods and Supplementary Materials**). Conventional clinical variables were transformed or constructed with the emphasis to be able to capture clinically relevant aspects of disease risk. For instance, we dichotomized continuous variables according to generally accepted clinical thresholds, thus obtaining for each of these clinical variables an ‘at risk’ [TRUE/FALSE] variable. For the same purpose, some variables were either merged or split. For instance, a sex-specific ‘*obesity*’ [TRUE/FALSE] variable was defined using body mass index, waist circumference and sex, whereas chronological age was split into three categories **(Figure 1B)**. Overall, we were able to construct and evaluate 20 variables mainly representing risk factors of cardio-metabolic health that are routinely assessed in epidemiological and clinical studies.

**Figure 1:**
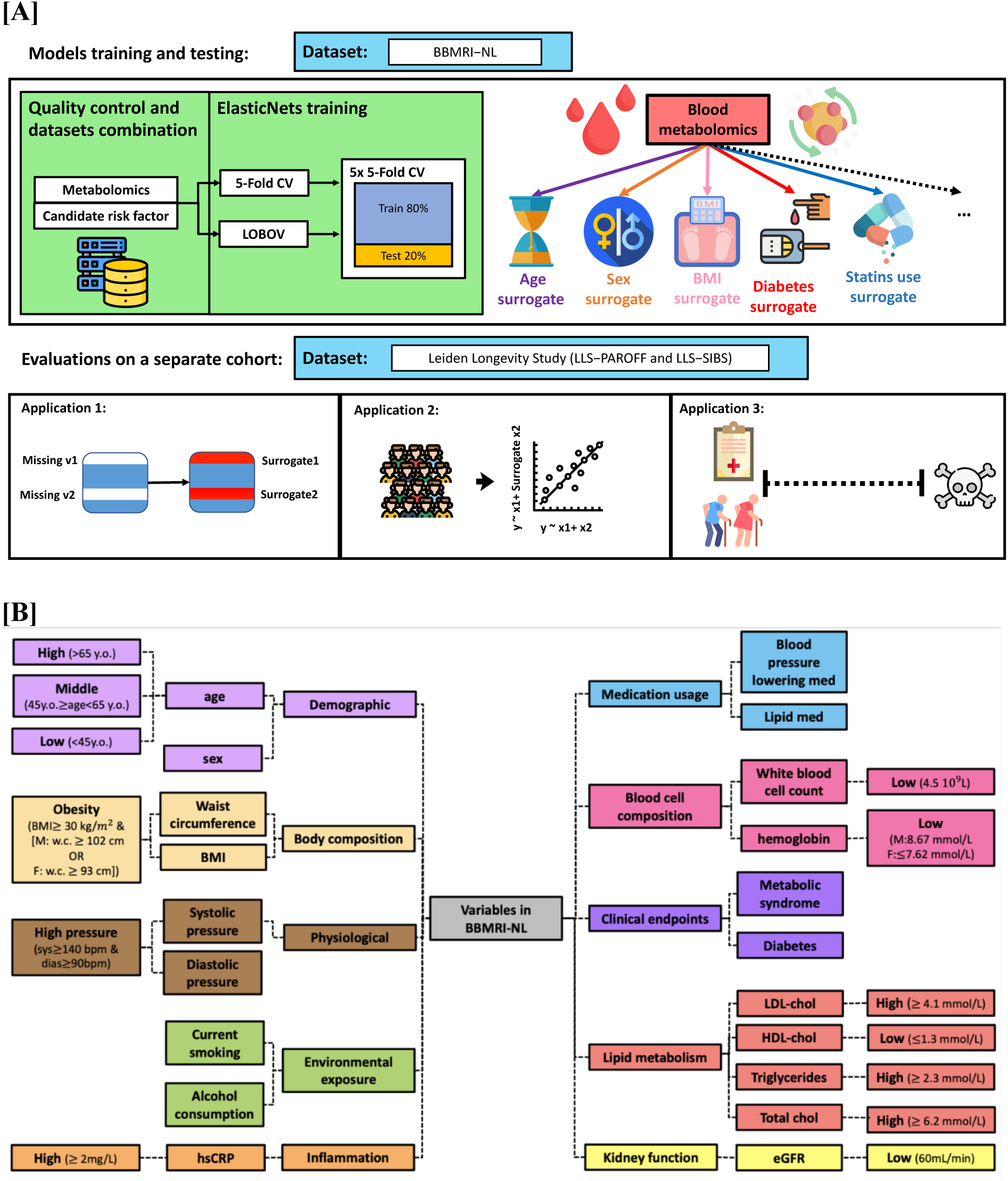
Study design. **[A]** Upper panel: Training of ^1^H-NMR metabolomics-based predictors for routinely assessed phenotypic variables available in BBMRI.nl. This data set was created as a collaboration of 28 community and hospital-based cohorts that collected nuclear magnetic resonance (^1^H-NMR) metabolomics data (Nightingale) for ∼31,000 individuals. Upper panel left: Metabolomics-based predictors were trained using an *inner* loop of 5-fold Cross Validation (CV) (with 5 repetitions) for hyperparameter optimization and were evaluated in unseen data employing an *outer* loop of 5-fold CV or Leave-One-Biobank-Out-Validation (LOBOV). Upper panel right: using our models 19 different surrogate values can be derived from a single metabolomics data measurement to impute or complement a broad set of conventional clinical variables routinely assessed in epidemiological and clinical studies. Lower panel: Trained metabolomics-predictors were evaluated in two application scenarios using a held-out study, the Leiden Longevity Study [20]. This study is a two-generation family-based cohort consisting of highly aged parents (LLS-SIBS, N = 817, median age = 92 years) and their middle-aged offspring and the partners thereof (LLS-PAROFF, N = 2,280, median age = 59 years), for which we had access to additional detailed phenotypic information. Trained predictors were evaluated for their ability to reconstruct missing datapoints in an independent dataset (Application 1, lower left), to be used as confounder in Metabolome Wide Association Studies (Application 2, lower central), and to investigate and to explore determinants of health in older individuals (Application 3, lower right). **[B]** Groupings of phenotypic variables routinely assessed in epidemiological and clinical studies for which data was available in BBMRI-NL. Continuous variables are dichotomized at levels generally accepted to confer an increased risk for cardio-metabolic endpoints. As various cutoffs on chronological age are in use, in part reflecting the highly non-linear relation between chronological age and disease risk, we choose to split chronological age in three categories (I ‘young’: < 45 years [TRUE/FALSE]; II ‘middle-aged’: ≥ 45 years [TRUE/FALSE]] and III ‘old’: < 65 years [TRUE/FALSE]]; ≥ 65 years). We integrated Body Mass Index, waist circumference and sex into one sex-specific measure of ‘obesity’. Similarly, we integrated diastolic blood pressure (DBP) and systolic blood pressure to arrive at one variable ‘high pressure’. Overall, we obtain data for 20 dichotomous phenotypic variables. Colors indicate groupings. Through the paper, we indicated some of the clinical variables with the following abbreviation: BMI=Body Mass Index, med=medication, e.g.: lipid or blood pressure lowering medication, hsCRP=high-sensitivity C-Reactive Protein, eGFR=estimated Glomerular Filtration Rate, chol=cholesterol, hgb=haemoglobin, wbc=white blood cells.

Logistic Elastic-NET regression models were trained for each of the 20 dichotomous variables, measured in at least 6 of the BBMRI studies, in both healthy and diseased individuals. Model development was performed in two loops to prevent overtraining. An *inner* loop of 5-Fold Cross Validation with 5 repetitions was used to tune the hyperparameters of the model. Model performances where then evaluated in an *outer* loop of held out data, using again a 5-Fold CV or a Leave-One-Biobank-Out-Validation (LOBOV) (**Figure 1A, Methods**). We assessed model performances using the mean Area Under the Curve (AUC) of the receiver-operator curve obtained in the *outer* 5-Fold CV (**Table 1**) and considered a model’s performance to be sufficiently accurate at AUC > 0.7. Overall, 19 out of 20 models passed this criterium, with only a single phenotypic variable, ‘*high-pressure*’, that could not be accurately captured by ^1^H-NMR (AUC_5-Fold CV_ = 0.68). Strikingly, 9 out of 20 models achieved an AUC_5-Fold CV_ > 0.9. While some of these high performances are expected as they directly relate to metabolic markers assessed on the platform (‘*Low eGFR*’, ‘*high triglycerides*’, ‘*high LDL cholesterol*’, ‘*high total cholesterol*’, and ‘*low LDL cholesterol*’), this is not the case for four other high performing models: ‘*diabetes*’ (AUC_5-Fold CV_ = 0.94), ‘*metabolic syndrome*’ (AUC_5-Fold CV_ = 0.93), ‘*sex*’ (AUC_5-FoldCV_ = 0.92), ‘*lipid medication*’ (AUC_5-Fold CV_ = 0.90). Also, other important cardio-metabolic health statuses, including ‘*obesity*’, ‘*high CRP*’ and ‘*blood pressure lowering medication*’ were predicted at a more than satisfactory accuracy (AUC_5-Fold CV_ > 0.8), indicating that overall, the ^1^H-NMR metabolome can be used to impute a broad spectrum of common clinical variables.

**Table 1.**
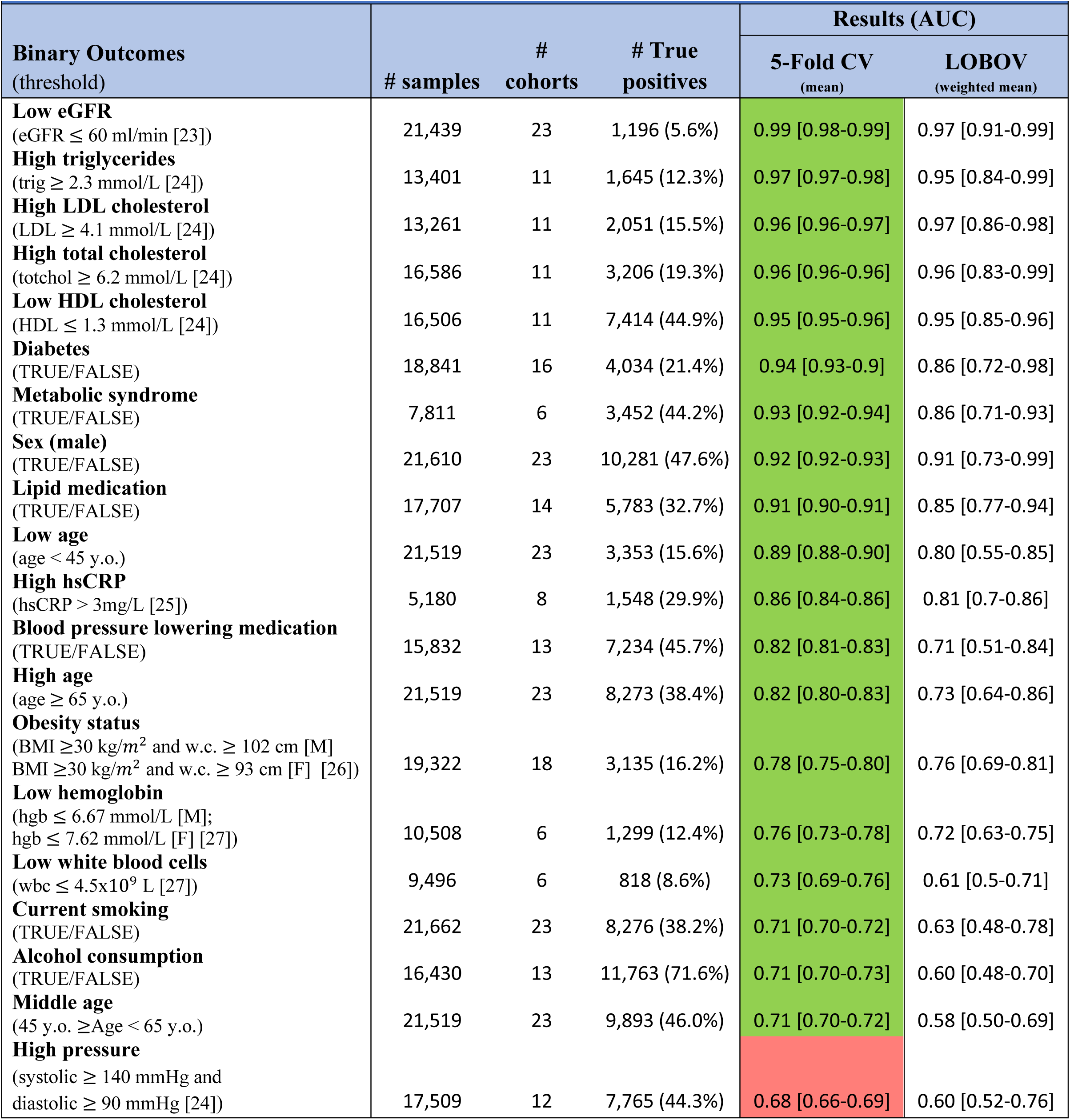
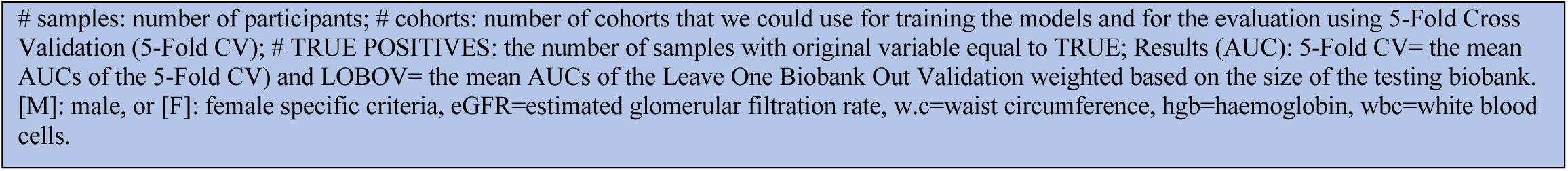
Performances of the 20 metabolic predictors

| Table 1 \| Performances of the 20 metabolic predictors |  |  |  |  |  |
| --- | --- | --- | --- | --- | --- |
| Binary Outcomes<br>(threshold) | # samples | # cohorts | # True positives | Results (AUC) |  |
|  |  |  |  | 5-Fold CV<br>(mean) | LOBOV<br>(weighted mean) |
| **Low eGFR**<br>(eGFR $\leq 60$ ml/min [23]) | 21,439 | 23 | 1,196 (5.6%) | 0.99 [0.98-0.99] | 0.97 [0.91-0.99] |
| **High triglycerides**<br>(trig $\geq 2.3$ mmol/L [24]) | 13,401 | 11 | 1,645 (12.3%) | 0.97 [0.97-0.98] | 0.95 [0.84-0.99] |
| **High LDL cholesterol**<br>(LDL $\geq 4.1$ mmol/L [24]) | 13,261 | 11 | 2,051 (15.5%) | 0.96 [0.96-0.97] | 0.97 [0.86-0.98] |
| **High total cholesterol**<br>(totchol $\geq 6.2$ mmol/L [24]) | 16,586 | 11 | 3,206 (19.3%) | 0.96 [0.96-0.96] | 0.96 [0.83-0.99] |
| **Low HDL cholesterol**<br>(HDL $\leq 1.3$ mmol/L [24]) | 16,506 | 11 | 7,414 (44.9%) | 0.95 [0.95-0.96] | 0.95 [0.85-0.96] |
| **Diabetes**<br>(TRUE/FALSE) | 18,841 | 16 | 4,034 (21.4%) | 0.94 [0.93-0.9] | 0.86 [0.72-0.98] |
| **Metabolic syndrome**<br>(TRUE/FALSE) | 7,811 | 6 | 3,452 (44.2%) | 0.93 [0.92-0.94] | 0.86 [0.71-0.93] |
| **Sex (male)**<br>(TRUE/FALSE) | 21,610 | 23 | 10,281 (47.6%) | 0.92 [0.92-0.93] | 0.91 [0.73-0.99] |
| **Lipid medication**<br>(TRUE/FALSE) | 17,707 | 14 | 5,783 (32.7%) | 0.91 [0.90-0.91] | 0.85 [0.77-0.94] |
| **Low age**<br>(age $< 45$ y.o.) | 21,519 | 23 | 3,353 (15.6%) | 0.89 [0.88-0.90] | 0.80 [0.55-0.85] |
| **High hsCRP**<br>(hsCRP $> 3$mg/L [25]) | 5,180 | 8 | 1,548 (29.9%) | 0.86 [0.84-0.86] | 0.81 [0.7-0.86] |
| **Blood pressure lowering medication**<br>(TRUE/FALSE) | 15,832 | 13 | 7,234 (45.7%) | 0.82 [0.81-0.83] | 0.71 [0.51-0.84] |
| **High age**<br>(age $\geq 65$ y.o.) | 21,519 | 23 | 8,273 (38.4%) | 0.82 [0.80-0.83] | 0.73 [0.64-0.86] |
| **Obesity status**<br>(BMI $\geq 30$ kg/m<sup>2</sup> and w.c. $\geq 102$ cm [M]<br>BMI $\geq 30$ kg/m<sup>2</sup> and w.c. $\geq 93$ cm [F] [26]) | 19,322 | 18 | 3,135 (16.2%) | 0.78 [0.75-0.80] | 0.76 [0.69-0.81] |
| **Low hemoglobin**<br>(hgb $\leq 6.67$ mmol/L [M];<br>hgb $\leq 7.62$ mmol/L [F] [27]) | 10,508 | 6 | 1,299 (12.4%) | 0.76 [0.73-0.78] | 0.72 [0.63-0.75] |
| **Low white blood cells**<br>(wbc $\leq 4.5 \times 10^9$ L [27]) | 9,496 | 6 | 818 (8.6%) | 0.73 [0.69-0.76] | 0.61 [0.5-0.71] |
| **Current smoking**<br>(TRUE/FALSE) | 21,662 | 23 | 8,276 (38.2%) | 0.71 [0.70-0.72] | 0.63 [0.48-0.78] |
| **Alcohol consumption**<br>(TRUE/FALSE) | 16,430 | 13 | 11,763 (71.6%) | 0.71 [0.70-0.73] | 0.60 [0.48-0.70] |
| **Middle age**<br>(45 y.o. ≥ Age $< 65$ y.o.) | 21,519 | 23 | 9,893 (46.0%) | 0.71 [0.70-0.72] | 0.58 [0.50-0.69] |
| **High pressure**<br>(systolic $\geq 140$ mmHg and<br>diastolic $\geq 90$ mmHg [24]) | 17,509 | 12 | 7,765 (44.3%) | 0.68 [0.66-0.69] | 0.60 [0.52-0.76] |

As the performances of our models may vary per biobank due to study-specific characteristics, e.g. varying study inclusion criteria or protocols for sample storage, we also evaluated the variation of our model performances across biobanks. First, using a Leave-One-Biobank-Out-Validation (LOBOV), we evaluate how our models would perform when applied to data of a new unseen biobank. As expected, mean model accuracies of the LOBOV, weighted based on the size of the testing biobank, show more variation across folds (**Figure S2A-B)** and are generally slightly lower than the overall results of the 5-Fold CV (**Table 1**). In particular, some of the smaller studies containing diseased patients showed relatively poor accuracies (**Figure S2B**). Indeed, surrogate values do show cohort specific effects, but interestingly, this does not necessarily affect its predictive performance within cohorts **(Figure S2C)**. Overall, 14 out of the 20 models performed on average satisfactorily (AUC_LOBOV_ > 0.7) across all studies in the LOBOV setting.

### Metabolic surrogates show dependencies mimicking the conventional clinical variables

Given that all models are trained on a relatively limited set of metabolic markers, we investigated to what extent the produced models and predictions show mutual dependencies. For this purpose, we first visualized the coefficients (betas) of the logistic Elastic-NETs (**Figure 2**) to show the relative importance of the metabolites within each of the prediction models. While the selection of variables for the models shows a distinct pattern, we also note some similarities, as quantified by the correlations between the model coefficients (**Figure S3**). Overall, we note a clear preference for the models to include metabolites of the classes “Lipoproteins” and “Lipids and related measures” over “Amino Acids”. In addition, we note that the models of related phenotypes also display some resemblances in the employed features, for instance ‘*lipid medication*’ and ‘*blood_pressure_lowering_medication’* share some model characteristics.

**Figure 2:**
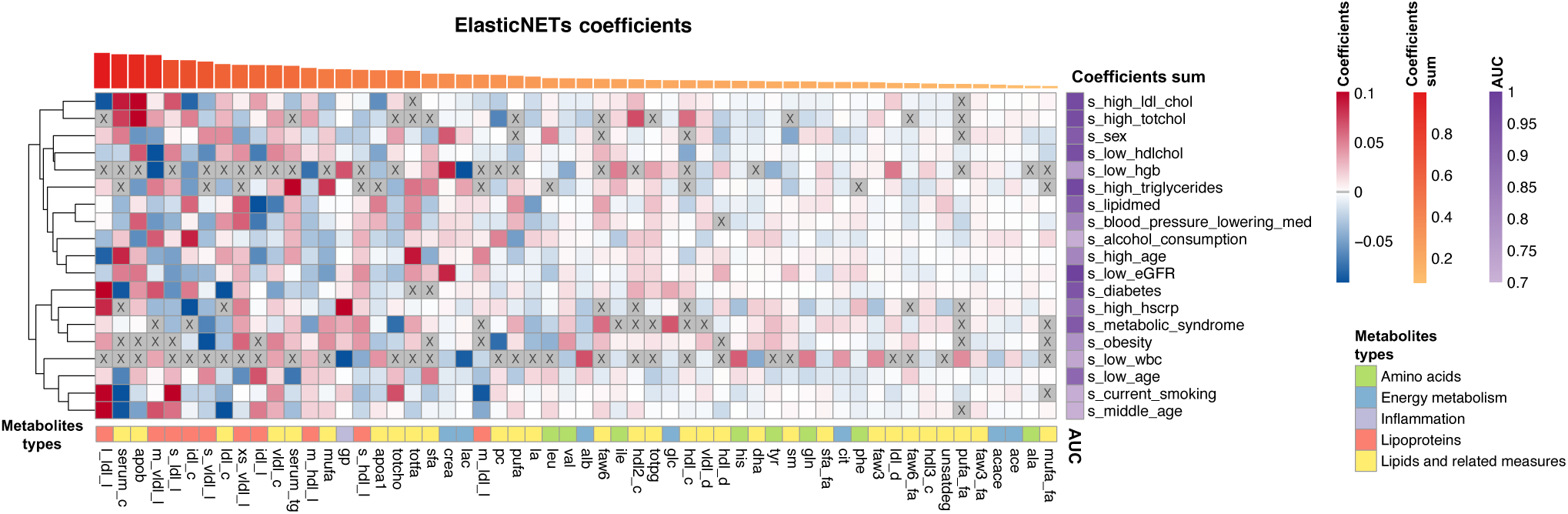
ElasticNETs metabolites relative importance. The heatmap reports the relative importance of the metabolites (columns) in each of the trained models (rows). Prior to visualization, metabolite coefficients were scaled per model by dividing them by the coefficients’ sum in each model to create the relative importance per model. Top: Metabolites were then ordered based on the sum of their importance across all models. In addition, the models are clustered on the similarity between relative importance. Bottom: Categorized metabolic measures: “Amino acids”, “energy metabolism”, “inflammation”, “lipoproteins” and “lipids and related measures”. Right: Mean AUCs of the 5-FoldCV in a scale of purple.

We next evaluated correlations between the outputs of our models, from here on referred to as the ‘metabolic surrogates’ (**Figure 3)** and compared these to correlations between the original clinical variables (**Figure S1B**) in the BBMRI.nl data set. Overall, we observe that the model outputs show correlation patterns and groupings that largely mimic that of the original variables. For instance, model outputs trained on variables related to weight problems, i.e. ‘*obesity*’, ‘*diabetes*’, ‘*metabolic syndrome*’, show high mutual correlations, and moreover are grouped with model outputs trained on medication usage, i.e. ‘*lipid medication*’ and ‘*blood pressure lowering medication*’. Although we observe some correlations between the outputs of our different age predictors i.e. ‘*low age*’, ‘*middle age*’, ‘*high age*’, we observe that ‘*high age*’ is grouped with the models for ‘*high hscrp’*, ‘*lipid medication*’ and ‘*blood pressure lowering medication*’, while ‘*middle age*’ is grouped with ‘*current smoking*’ and ‘*alcohol use*’ and ‘*low age*’ with ‘*low white blood cell count’*. This suggests that at different ages, different conventional clinical variables play a role in physiology; an aspect well-known from literature [28]–[30]. Overall, this indicates that our models show mutual dependencies similar as we observe for the original clinical variables.

**Figure 3:**
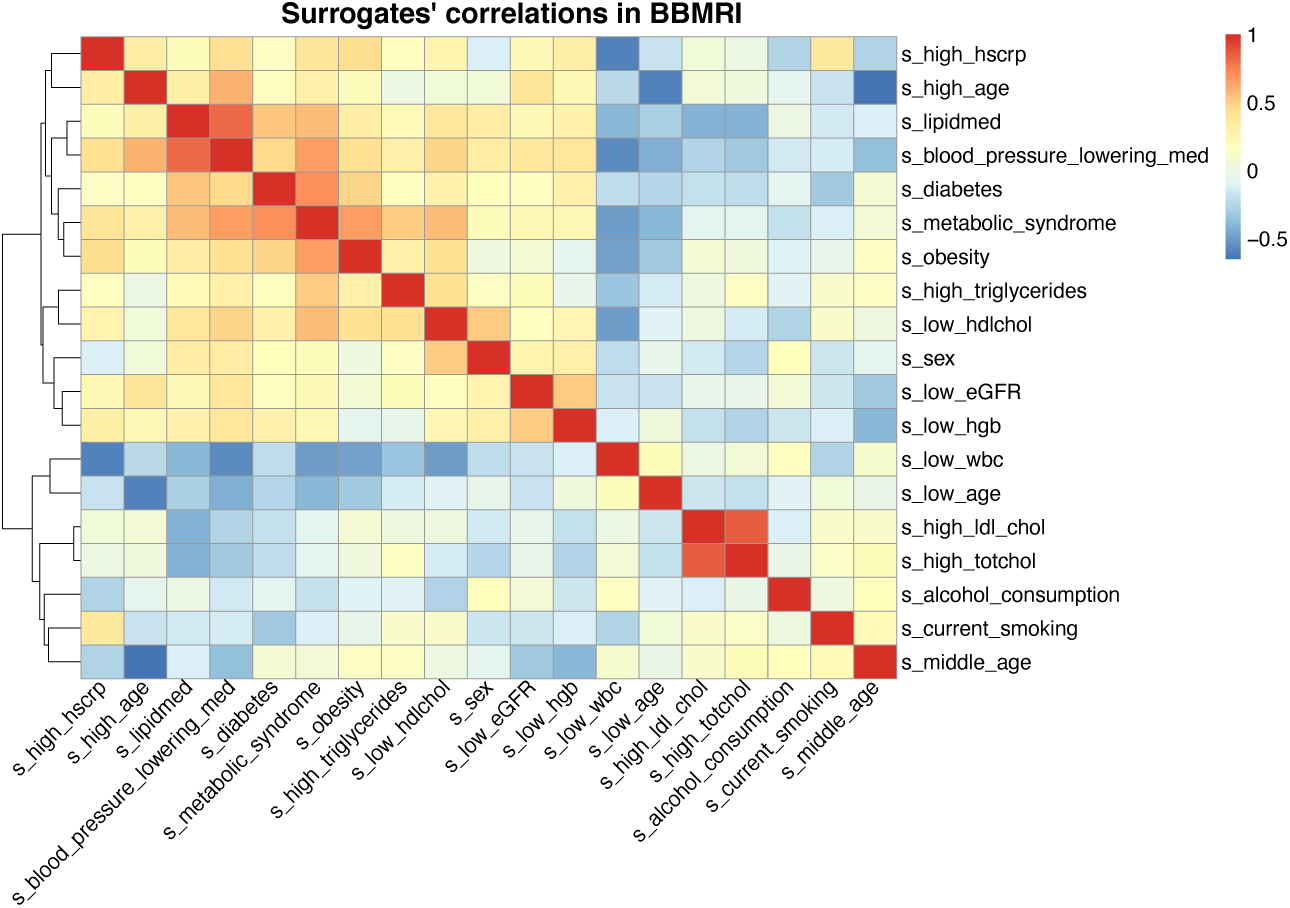
Heatmap of pairwise correlations of the metabolic based surrogate markers calculated in BBMRI-NL. The heatmap of correlations of the metabolic surrogate values of the 19 successful models, clustered based on the correlation levels, between the imputed metabolic surrogate levels within BBMRI-NL.

### Projection in an independent study demonstrates model accuracy

We performed a more extensive evaluation of the surrogate values by employing Nightingale ^1^H-NMR metabolomics and phenotypic data of the Leiden Longevity Study, a cohort excluded from the training and testing sets [20]. The Leiden Longevity Study is a two-generation family-based cohort consisting of highly aged parents (LLS-SIBS, 851 individuals, age median = 92 years old) their middle-aged offspring and their partners (LLS-PAROFF, 2,307 individuals, age median = 59 years old). Using our models to project metabolic surrogates in the LLS-PAROFF gave an independent confirmation that conventional clinical variables can be readily captured by ^1^H-NMR metabolomics. Splitting the surrogate values by the actual labels of the corresponding binary phenotypes generally showed a good separation for important cardio-metabolic variables like ‘*sex*’, ‘*diabetes status*’, ‘*lipid medication’, ‘blood_pressure_lowering_medication’* and ‘*high LDL cholesterol*’ (**Figure 4A** and **S4**), emphasizing the suitability of our models for quality control purposes or to impute missing data. For instance, model results for ‘sex’ could be applied to verify absence of sample mix-ups (*t*. *stat* = 44.58, *p* = 1.4*x*10^-313^). In addition, surrogate values seem informative on the nature of the missingness of phenotypic data. For instance, participants with a missing diabetes status typically had metabolic surrogate values similar to those of participants without diabetes (diabetes: *μ_F_*= 0.05, *μ_T_* = 0.41, *μ_NA_* = 0.08), suggesting that a missing diabetes status generally implies ‘non-diabetics’ in this cohort. Similar observations were made for medication status (lipidmed: *μ_F_* = 0.22, *μ_T_* = 0.45, *μ_NA_* = 0.21 and blood_pressure_lowering_med: *μ_F_*= 0.44, *μ_T_* = 0.55, *μ_NA_* = 0.46): participants with missing statuses were more similar to non-medication users than medication users. In contrast, participants with missing values in LDL cholesterol had surrogate values indicating “at risk” levels of LDL cholesterol (high_ldl_chol: *μ_F_*= 0.07, *μ_T_* = 0.66, *μ_NA_* = 0.14). Lastly, our surrogates also allow for explorative analyses of totally unrecorded variables. For instance, the ‘*metabolic syndrome*’ surrogate indicates participants who are more likely to have metabolic syndrome, a status which was not assessed in the LLS-PAROFF cohort (**Figure 4A)**.

**Figure 4:**
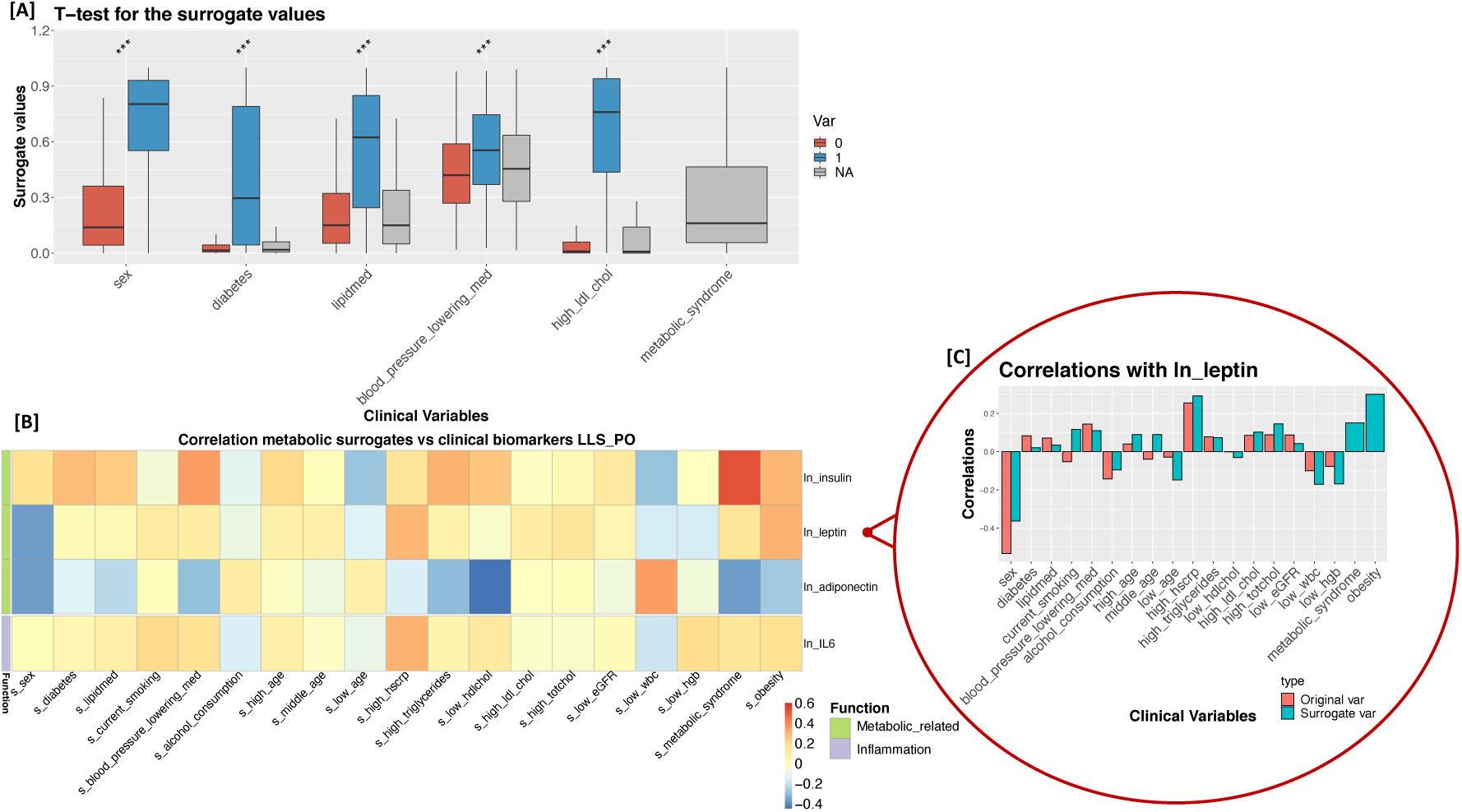
Metabolic surrogates applied to LLS-PAROFF: **[A]** Paired boxplots show surrogate values split between the TRUE/FALSE (0/1) in the original values of the clinical variables (*** *p* value ≤ 0.001). For metabolic syndrome the original variables are entirely missing, so no *p*-value is reported. **[B]** Heatmap of the correlations between the metabolomic surrogates (columns) and four additional cardio-metabolic biomarkers (rows) available in LLS-PAROFF. Values of insulin, leptin, adiponectin and IL6 were transformed with a natural logarithm. **[C]** Paired bar plots comparing the correlations computed between leptin and the metabolic surrogates (blue) and those computed between leptin and the values of the original clinical variables (red) the surrogates are trained to predict.

### Projection in the Leiden Longevity Study shows associations with additional cardio-metabolic phenotypes

Within the LLS-PAROFF we had access to several additional variables pertaining to one’s cardio-metabolic risk namely hormone levels of insulin, leptin, and adiponectin, as well as the levels of the inflammatory marker interleukin 6 (*IL6*) (**Figure 4B**). As expected, insulin levels correlated positively with most surrogate cardio-metabolic risk factors and endpoints, including ‘*diabetes*’ [31] (*r* = 0.28, *p* = 7.6*x*10^-42^) and even more so with *‘metabolic syndrome’* (*r* = 0.52, *p* = 1.4*x*10^-152^) [32]. Conversely, both ‘*low wbc*’ and ‘*low age’* were inversely correlated with insulin, both reflecting the associations with decreased insulin sensitivity in those with high white blood cell counts [33] or in old age [34]. A similar analysis for the satiety hormone leptin showed the strongest positive correlations with *‘obesity’* [35] (*r* = 0.3, *p* = 5.6*x*10^-49^), but also with ‘*high hscrp*’[36] (*r* = 0.29, *p* = 4.2*x*10^-46^). A significant correlation with leptin was also found for *‘metabolic syndrome’* [37] (*r* = 0.15, *p* = 3.9*x*10^-13^), yet not ‘*diabetes*’ [38] (*r* = 0.02, *p* = 0.33). In line with previous studies, higher levels of the adiponectin hormone generally correlated with lower values of the surrogates, most prominently with ‘*low hdlchol*’ ( *r* = −0.47, *p* = 8.6*x*10^-123^) and ‘*high triglycerides*’ (*r* = 0.3, *p* = 1.1*x*10^-48^) [39]. Higher adiponectin levels were positively correlated with ‘*low wbc*’(*r* = 0.35, *p* = 6.1*x*10^-68^), reproducing the previously reported association by Matsubara *et al* [40]. Levels of the inflammatory marker IL6 were most prominently positively correlated with the surrogates ‘*high hscrp’* [41] (*r* = 0.31, *p* = 8.7*x*10^-51^), *‘current smoking’* [42] (*r* = 0.2, *p* = 3.9*x*10^-13^), and inversely correlated with ‘*low wbc*’ [43] (*r* = −0.19, *p* = 3.9*x*10^-13^). When comparing these correlation patterns obtained with the surrogate values (**Figure 4B**) with those you would get when using the original values (**Figure S7A**), we generally notice highly similar trends (*r* ∼ 0.83, **Figure S7F**), as exemplified for leptin (**Figure 4C**). Overall, these findings indicate that our metabolic surrogates faithfully reproduce the original clinical variables in their association with insulin, leptin, adiponectin and IL6.

### Metabolic surrogates to explore confounders in Metabolome Wide Association Studies

We next explored the use of ^1^H-NMR metabolic surrogates to complement missing phenotypic data in metabolome-wide association studies (MetaboWAS). As an example, we evaluated the association of metabolic markers with Type 2 Diabetes status (T2D) in absence of information on a known potential confounder: BMI. We designed a controlled experiment to evaluate to what extent surrogate ‘obesity’ can replace BMI, using data of 1,697 individuals of LLS-PAROFF with complete metabolomic, BMI and diabetes status, of which 79 are diagnosed with type 2 diabetes.

First, we ascertained that BMI was indeed a confounder, also within the LLS-PAROFF, by showing that BMI associated with the outcome (Type 2 Diabetes status, *t*-*test*= −7.83, *p* = 8.25×10^-15^ **Figure S8A**), as well as many of the determinants (147 significant metabolites after correction, see **Methods**) of the MetaboWAS. Concomitantly, further adjustment of the MetaboWAS on T2D for BMI drastically reduced the number of significant metabolites. To compare, when adjusting for age and sex we identified 136 metabolites significantly associated with diabetes status, whereas further adjustment for BMI identified 80 significant metabolites (**Figure S9B Comparison 1**). Next, we performed the same association analyses using the ‘*obesity*’ surrogate as confounder. Similar to BMI, also the ‘*obesity*’ surrogate is significantly higher in diabetics as compared to non-diabetics (*t*-*test* = −11.2, *p* = 2.48×10^-28^ **Figure S8B**) and was associated with many of the metabolites (176 significant associations). Further adjusting the MetaboWAS on T2D for ‘*obesity*’ reduced the number of significant metabolites to 66 (**Figure S9B Comparison 2**).

We then investigated to what extent adjusting for BMI or adjusting for the ‘*obesity*’ surrogate yields similar metabolite markers to T2D associations, by comparing the obtained estimates from both models (**Figure 5**). Overall, highly similar (*r*^2^ = 0.902) associations between metabolic markers and T2D are found for both models, with glucose being the most significantly associated marker in both (*p* = 9.43*x*10^-28^ when correcting for BMI and *p* = 1.6*x*10^-26^ correcting for *‘obesity’*). While most metabolites reported to be significantly associated with T2D overlap between the two models (62 out of 227; in purple), some discrepancies were observed, particularly at the significance threshold. When correcting for BMI, 18 significant metabolic markers were identified, that were not identified when correcting for ‘*obesity*’ (red dots, false negative rate ∼ 0.11). Conversely, 7 metabolites were deemed significantly associated with diabetes status when adjusting for ‘*obesity*’, but not when adjusting for BMI (blue dots, false positive rate ∼ 0.027). Nevertheless, overall, the differences in estimated effects remain small, indicating that metabolic surrogates may prove useful to account for missing data in epidemiological studies.

**Figure 5:**
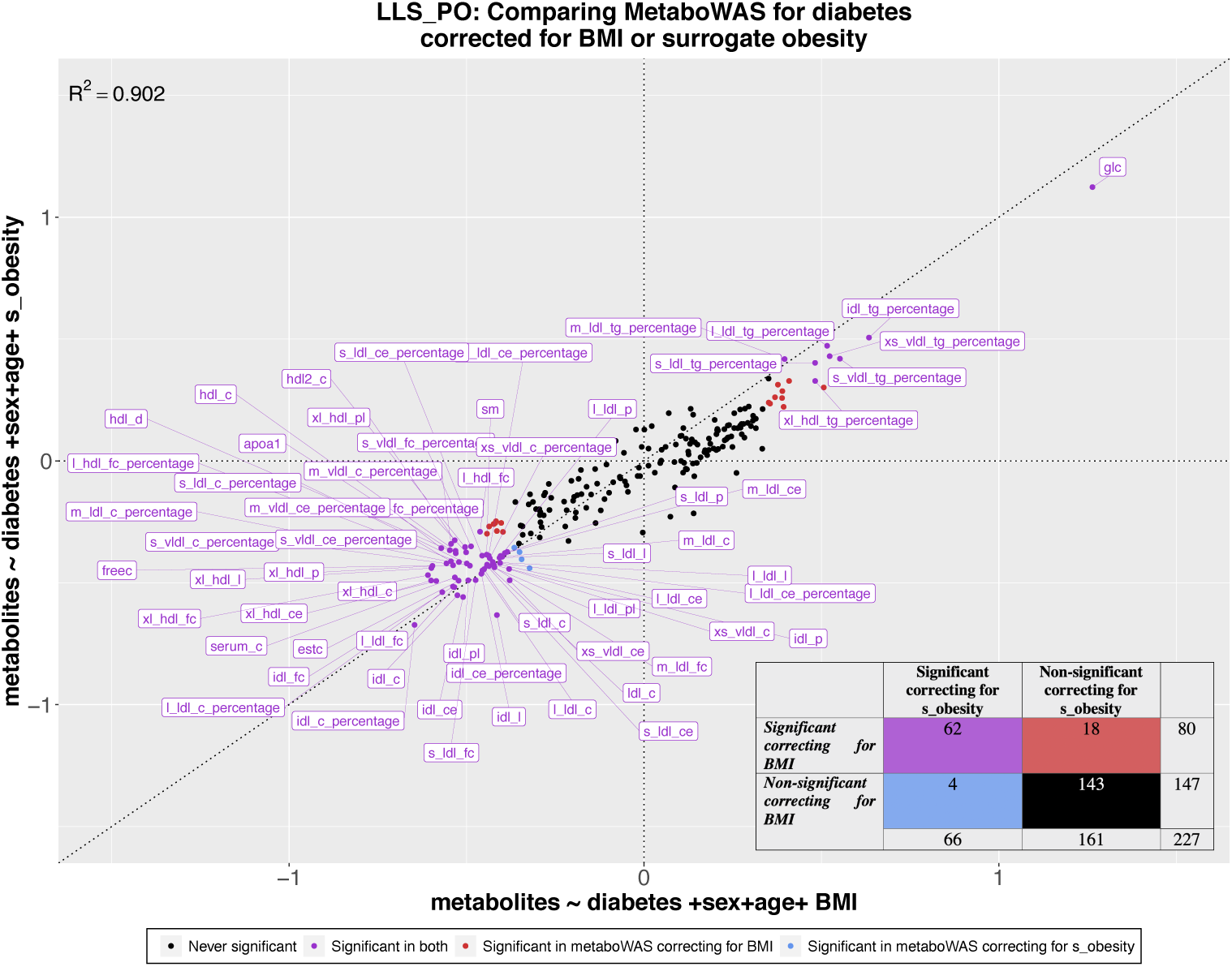
Comparison between the estimated coefficients of metaboWAS on T2D adjusted for BMI or for surrogate *‘obesity’*. On the x-axis the metaboWAS for diabetes adjusting for BMI and on the y-axis the metaboWAS for diabetes adjusted for surrogate obesity. The data set composed of 1,697 individuals, 79 of which are diabetics. Estimated coefficients for each metabolite (points) are colored based on their significance in the two models: purple: significant in both; red: significant when adjusted for BMI only; blue: significant when adjusted for surrogate obesity only; black never significant. Lower right corner: a contingency table with the number of significant and non-significant metabolites identified using the two models.

### Metabolic surrogates associate with incident all-cause mortality in older individuals

Next, we evaluated whether metabolic surrogates are indicative of health at old age, by associating these with all-cause mortality in a nonagenarian subsample of the Leiden Longevity Study (LLS_SIBS; 844 individuals, median age at baseline: 92 years old) (**Figure 6A**). Using a Cox proportional hazards model adjusted for sex and age at inclusion for each of the 19 metabolic surrogates (**Materials and Methods**), we observed that 13 out of the 19 surrogates associated significantly with all-cause mortality (**Figure 6,** ‘all’). In line with previous reports, we observed the largest effect sizes with the surrogate levels of ‘*high age’*, ‘*medications usage’*, ‘*diabetes status’,* ‘*high hscrp’* and ‘*hemoglobin’.* As previous studies have reported sex-specific associations for these clinical variables with all-cause mortality, we conducted a stratified analysis [44]–[50]. Although, the direction of association with all-cause mortality remains generally the same between men and women, the strengths and their significance are in some cases different. For instance, the surrogate ‘*diabetes’* is associated with a higher risk on mortality in men (HR = 1.23, *p* = 6.42×10^-4^), than for women (HR = 1.11, *p* = 3.1×10^-3^), the same goes for ‘*blood pressure lowering medication’* (men: HR = 1.3, *p* = 1.13×10^-5^, women: HR = 1.1, *p* = 6.39×10^-3^). In contrast, ‘*low hemoglobin*’ is associated with a higher risk in women (HR = 1.5, *p* = 1.95×10^-13^), than men (HR = 1.37, FDR = 2.42×10^-8^).

**Figure 6:**
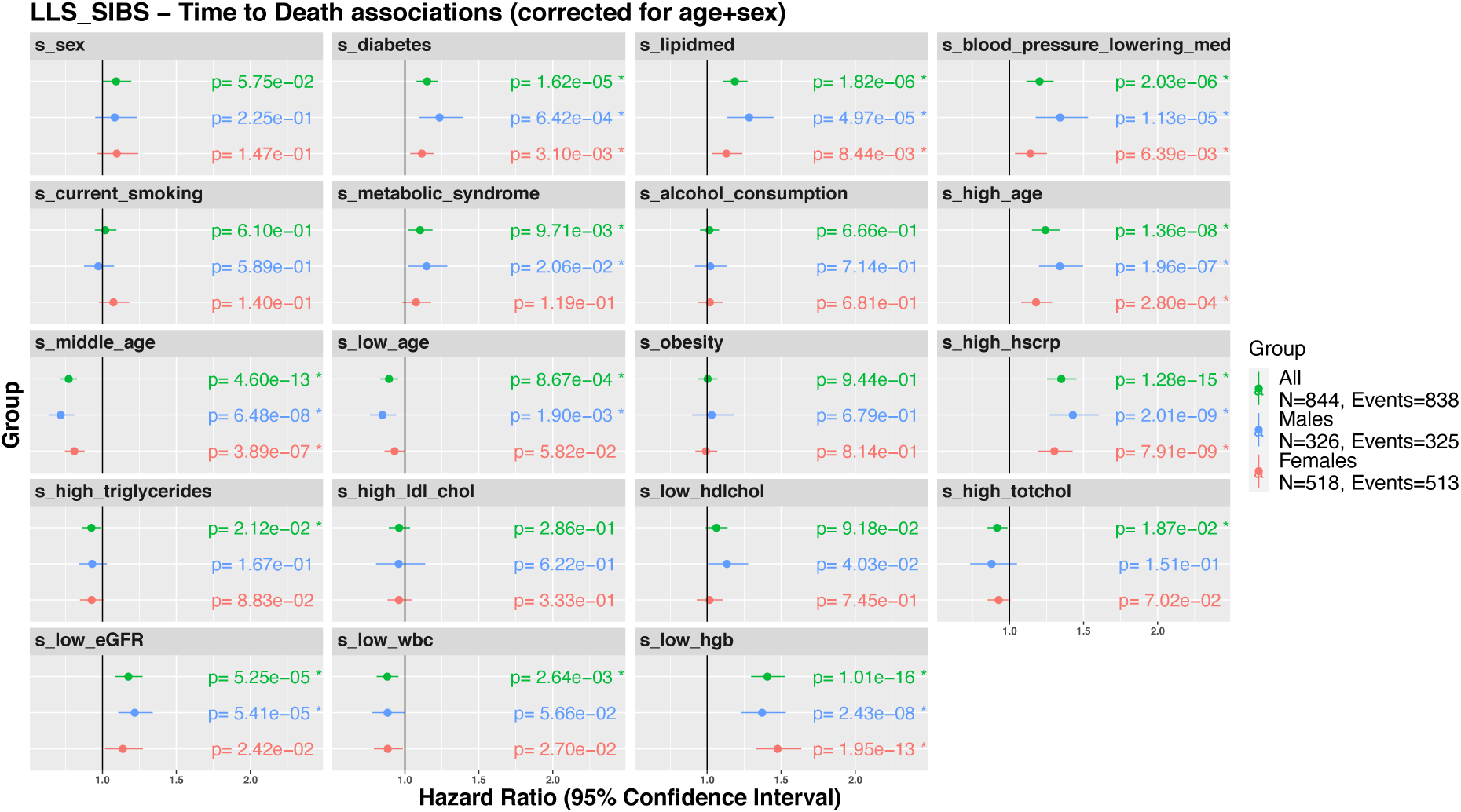
Associations of the surrogate metabolic measures with incident all-cause mortality. Associations of the metabolic surrogates with time to all-cause mortality in LLS-SIBS, in groups comprising the entire set (“All”, N = 844 with 838 reported deaths), males (N = 326 with 325 reported deaths) or females (N = 518 with 513 reported deaths).

To identify the minimal set of metabolic surrogates independently associating with all-cause mortality, we performed a stepwise (forward/backward) cox regression, adjusted for age at sampling, in the LLS-SIBS dataset (**Figure 7A-B**), stratified for sex. The surrogates ‘*high hsCRP*’ and ‘*high triglycerides*’ emerged as independent predictive features in both male and female models, associated with an increased and decreased risk respectively. While eight surrogates contributed to the male model, including ‘*lipid medication’*, ‘*high age*’, ‘*high hsCRP’* and ‘*low hdlchol’*, only three surrogates contributed to the mortality prediction in females: ‘*high hsCRP’*, ‘*high triglycerides’* and *‘low hemoglobin’*. These findings are in line with previous reports that different risk factors seem to predict survival up to the highest ages for the different sexes [51]–[53].

**Figure 7:**
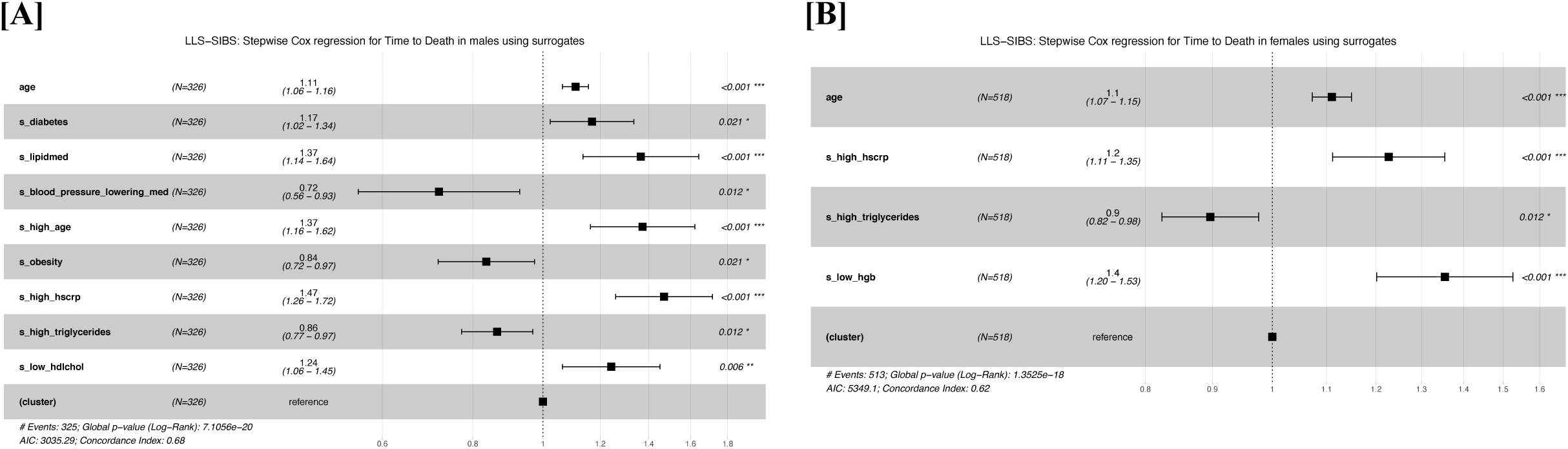
Composite metabolomics predictors of incident all-cause mortality: Predictors of time to death for males **[A]** and females **[B]**, in LLS-SIBS, composed using the surrogate metabolic measures, sex and age. “(cluster)” refers to the variable controlling for family relationships (methods). Cox regression models were made using a step forward/backward selection.

## Discussion

Missing phenotypic data is common in large epidemiological studies and in particular impedes biomarker research in older individuals. We employed ^1^H-NMR metabolomics data as a single source of information to successfully impute 19 out of 20 conventional clinical variables that mainly relate to cardio-metabolic health. We highlighted the potential of our imputation models for conventional clinical variables with three application scenarios. First, we applied our models to an independent study, the Leiden Longevity Study, demonstrating that we can reconstruct conventional clinical variables at high accuracy. Secondly, we showed the value of metabolic surrogates in omics studies when data on potential confounders is missing. Finally, we exemplified how metabolic surrogates can be used to explore risk factors of health in older individuals by showing that multiple metabolic surrogates are independently predictive of all-cause mortality.

Using logistic ElasticNET regression models we were able to reconstruct a broad range of conventional clinical variables assessed in BBMRI-NL pertaining to physiological measures, body composition measures, environmental exposures, inflammatory factors, medication usage blood cell composition, lipids metabolism, and also clinical endpoints. For this purpose, we constructed composite variables that may better capture particular aspects of health, for instance, our ‘*obesity*’ variable integrates body mass index, waist circumference and sex to create a sex-specific measure for overweight. In addition, we chose to construct binary representations of the continuous clinical variables for several reasons. First, we binarized continuous variables for a practical reason – to be able to judge all models on the same criteria. Secondly, predicting continuous variables using linear ElasticNET regression models emphasizes the prediction of the extremes of a phenotypic distribution, i.e. the model will fit the most atypical participants, whereas the current approach emphasizes to predict the commonly populated phenotypic range in which participants become at risk. Thirdly, our models output a posterior probability that indicates the likelihood (a continuous score) of a sample belonging to one of two labels, e.g. *obese*/*non-obese*. In effect, these posteriors reconstitute part of the information lost when dichotomizing continues variables, as exemplified by the observed correlation patterns between surrogates that mimic the correlation patterns between the original variables.

Our pre-trained models for conventional clinical variables allow for the imputation of missing datapoints in partially incomplete phenotypic variables, and moreover they offer the opportunity to explore associations with completely unobserved phenotypic variables. The latter is very much in line with the current use of PolyGenic Scores (PGSs) [54]–[56]. A PGS captures the genetic propensity of the realization of a particular polygenic phenotype. Nearly a thousand PGSs have been collected [57], which can be used to systematically explore correlations between a measured variable of interest and a wide array of phenotypes-by-proxy in genetic studies. We propose a similar use for metabolic surrogates in large metabolomics studies, yet with two noteworthy distinctions. Whereas PGSs can arguably be used to tease out causality in so-called Mendelian Randomization studies [58], metabolomic surrogates cannot. In contrast, while PGSs often only explain a very modest part of their respective phenotypes, metabolic surrogates explain a much larger part, thus enabling different types of applications. We illustrated this in our second application scenario where we showcased the use of surrogates to explore potential confounding by non-assessed phenotypic variables in omics studies. While use of actual phenotypic variables will always be preferred over metabolic surrogates, the availability of these metabolic surrogates can thus be used to direct replication efforts or to inform the design of new or follow-up studies.

Besides anthropometric measures and other physiological characteristics, the blood ^1^H-NMR metabolome was previously also shown to capture aspects directly pertaining to health outcomes. In particular, we and others have previously reported ^1^H-NMR metabolomics-based risk estimators of cardiometabolic disease [59], [60], pneumonia and COVID infection [61], and all-cause mortality [17]. While this clearly illustrates the vast potential of the blood ^1^H-NMR metabolome as a universal readout for health outcomes, it also raises the question what factors give rise to metabolomic profiles associated with adverse outcomes. Given that the find variation in the ^1^H-NMR metabolome is the result of a complex interplay of both environmental and genetic factors, we evaluated whether our surrogates might give us a first indication. To do so, we tested which of our surrogates might be indicative of all-cause mortality in an elderly subset of the LLS-study. Intriguingly, by employing our pre-computed models as well as when we built multi-variate cox-regression models for time-to-death, we find metabolic surrogates that relate to conventional clinical risk factors known to associate with mortality risk at old age. Moreover, sex-stratified analyses recapitulate some of the known differences in mortality associations observed at old age, with for instance many more risk factors independently associated for mortality in males, as compared to females. These results illustrate that metabolic surrogates can aid in the interpretation of metabolomics-based risk estimators.

This study has several limitations. LOBOV analyses revealed that the trained surrogates may show study-specific effects that may relate to employed procedures of data collection or sample storage of the cohorts under investigation. While these artifacts may be addressed using batch-correction algorithms [62], or employing deep learning models for the prediction tasks, we note that differences between studies may also be due to valid biological reasons, such as differences in inclusion criteria. Secondly, the number of biomarkers captured by the targeted NMR platform is small compared to the whole human metabolome (over 19,000 according to the Human Metabolome Database [63]). Therefore, more elaborate, though typically more costly, high-throughput platforms might reach even higher accuracy levels. However, employing more biomarkers also has the danger of overfitting to the aforementioned study-related artifacts.

In conclusion, we have shown that the blood metabolome assayed by ^1^H-NMR metabolomics can successfully capture a broad set of conventional clinical variables opening various possibilities to exploit surrogates of these clinical variables in in large epidemiological and clinical studies.

## MATERIALS and METHODS

### 1. Study populations

The samples used for the current study are part of the BBMRI-NL Consortium (Dutch Biobanking and BioMolecular resources and Research Infrastructure, https://www.bbmri.nl/), which includes the following 28 Dutch biobanks: ALPHAOMEGA, BIOMARCS, CHARM, CHECK, CODAM, CSF, DMS, DZS_WF, ERF, FUNCTGENOMICS, GARP, HELIUS, HOF, LIFELINES, LLS_PARTOFFS, LLS_SIBS, MRS, NESDA, PROSPER, RAAK, RS, STABILITEIT, STEMI_GIPS-III, TACTICS, TOMAAT, UCORBIO, VUMC_ADC, VUNTR. A description of the cohorts included is provided in the Supplementary Materials. Ethics committees approved the protocols for these studies in all the involved institutes, and all participants provided informed consent. The whole data set contains samples of ∼31,000 individuals.

### 2. Metabolomic measurements

The present study included metabolite concentrations measured in EDTA plasma samples using the high-throughput proton Nuclear Magnetic Resonance (^1^H-NMR) metabolomics (Brainshake Ltd./Nightingale Health^©^, Helsinki, Finland). This device provides the quantification of routine lipids, lipoprotein subclasses, fatty acid composition and various low-molecular weight metabolites including amino acids, ketone bodies and glycolysis-related metabolites in molar concentration units. Details about the methods and applications of the NMR platform have been provided previously [22], [64]. The total amount of metabolic variables reported is 226 for EDTA plasma samples, including the ratios and derived measurements, but only 63 of these were considered for the current study, to prevent overfitting [21], [59]. The list comprises the total lipid concentrations, fatty acids composition and low-molecular-weight metabolites including ketone bodies, glycolysis-related metabolites, amino-acids and metabolites related to immunity and fluid balance (See the Supplementary Materials for a full list).

### 3. Data Pre-processing

#### a. Pre-processing of metabolomics data

We included in our analyses all the cohorts reporting on all the 63 metabolic biomarkers, therefore we omitted CODAM (N = 254) and VUNTR (N = 3,896), which are missing acetoacetate and glutamine, respectively. We also decided to not consider the metabolites with low detection rates in more than one cohort (3-hydroxybutyrate) or which frequently failed to reach the minimum detection threshold (XL_VLDL_L, XXL_VLDL_L, L_VLDL_L, XL_HDL_L, L_HDL_L). We removed outlier samples with 1 or more missing metabolic measure (232 removed samples), 1 or more zeroes per sample (74 removed samples) and samples with any metabolite concentration level more than 5 standard deviations away from the overall mean per metabolomic variable (604 removed samples). The remaining 551 missing values in the dataset were imputed using the function nipals of the R package pcaMethods, and we z-scaled the metabolic measures across all samples to have comparable concentration levels between metabolites. The final data matrix comprised 26,107 samples across 56 metabolic variables. For more details, see Supplementary Materials. The number of samples used to train a predictor for a clinical variable depended on the number of samples missing this phenotypic information (Table 1). More information about the range of each phenotype within each biobank can be found in the Supplementary Materials.

#### **b.** Binarization of the clinical variables

To emphasize the relevant clinical conditions, we used clinical thresholds to obtain dichotomous variables out of the set of the available continuous risk factors, separating between “normal” and “at risk” levels for each risk factor (in ***Table1*** and in the Supplementary Document 3).

#### **c.** Composed clinical variables

We chose to include some composed clinical variables: 1) LDL cholesterol, which was calculated using the Friedewald equation [65] with the measured hdl cholesterol, triglycerides levels and total cholesterol; 2) eGFR (estimated Glomerular Filtration Rate), which is a measure for the kidney filtration rate of an individual, was calculated using the creatinine-based CKD-EPI equation [66]; 3) obesity, which is a binary variable describing if a person is clinically obese or not variable that uses BMI, waist circumference and sex based on the fniding of Flint et al. [26]; 4) high pressure, a binary variables which defines high blood pressure by using systolic and diastolic blood pressure [24]; 5) low_hgb (low hemoglobin), which is a binary variables describing ‘at risk’ levels of hemoglobin by using hemoglobin and sex [27].

### 4. Estimation of the metabolic surrogates

#### a. Method selection

The models considered for each Risk Factor are logistic regression models:

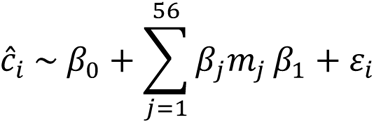

in which *c_i_*. represent one of clinical variables of interest, *m_j_* one of the 56 measured metabolites, *β_j_* the regression coefficient, and *ε_i_* the normal distributed reconstruction error. The regression coefficients are found by minimizing the ordinary least squares error, with the addition of an elastic net regularization term for the coefficients:

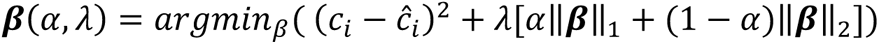

in which *λ* (∈ (0, ∞)) represents the “shrinkage parameter” and *α* (∈ (0, 1)) is the mixing parameter balancing the L1 and L2 norm regularizations. We fixed the mixing parameter *α* at 0.5 for the predictive models, like previously done by other authors and optimized the shrinkage parameter using an inner-fold cross-validation scheme.

#### **b.** Training and validation procedure

We employed two training-evaluation procedures to get an unbiased estimate of the models’ possible performances (Figure 1). As a first scenario, we used a Double 5-Fold-Cross-Validation (5-Fold CV) with 5 repetitions. This procedure consists of two loops of 5FCV, one internal and one external, in which we first split the dataset in testing (20%) and training (80%) sets and then on the latter set we have another 5FCV repeated for 5 different times, which is done for an unbiased tuning of the model (setting the correct *λ* parameter) that is finally trained on the complete training dataset and tested on the left-out test data. Both 5-FoldCVs were done such that the original distribution of each clinical variable is maintained as much as possible (using the function *createFolds* from the R package *caret*). In the second training-testing procedure, we applied a Leave-One-Biobank-Out-Validation (LOBOV), which consists of holding out one of the biobanks with the considered variable available, which is then used as a test set, while training on the remaining biobanks [16]. Also, in this setting, we applied a 5FCV with 5 repetitions to tune the best model for each training set.

### 5. Metabolome wide association studies

We conducted Metabolome Wide Association Studies (MetaboWAS) using the middle-aged cohort of the Leiden Longevity Study (LLS-PARTOFFs, 2,307 individuals, median age at baseline = 59 years old). As metabolites distributions are often skewed, we first transformed all metabolite measurements using a rank inverse normalization (RIN). Applying a PCA on the LLS-PARTOFFs dataset revealed that the first 40 principal components explain 99% of the variance in the metabolites (**Figure S9A**). Hence, the *p-value* of the MetaboWASes were Bonferroni corrected using 40 tests, i.e. a *p*-*value* designated significant when smaller than 0.00125 (0.05/40) [60]. We performed 5 different MetaboWASs.

### 6. Associations of the metabolic surrogates to all-cause mortality

We used Cox proportional hazards models with follow-up time as the time scale, to test for associations between the metabolic surrogate measures and incident endpoints, i.e. all-cause-mortality in LLS-SIBS. We checked for associations adjusting for age and sex. To avoid bias due to familial correlations from pedigrees, we used robust standard errors (calculated with the Huber sandwich estimator) implemented in R coxph function. Considering that the population in LLS-SIBS has a different inclusion criterium for men (age > 89 years old) and women (age > 91 years old), we also evaluated associations separately in men and women. *P-values* were corrected using Benjamini Hochberg separately for each selection (all individuals, men and women) and considered significant the FDR < 0.05. To select potentially interesting metabolic surrogate, we used a stepwise procedure for the Cox regression models, corrected for sex and age. Starting from a model containing the full set of available variables, we removed or added an unselected metabolic surrogate at each round based on the improvement on the model calculated from the Akaike Information Criterion and considering the *p*-value of each variable included in the model.

## Data Availability

Data available upon request at bbmri.nl

https://www.bbmri.nl/samples-images-data

## Acknowledgements

This work was performed within the framework of the BBMRI Metabolomics Consortium funded by BBMRI-NL (a research infrastructure financed by the Dutch government, NWO 184.021.007 and 184.033.111), by X-omics (NWO 184.034.019), VOILA (ZonMW 457001001) and Medical Delta (scientific program METABODELTA: Metabolomics for clinical advances in the Medical Delta). EvdA is funded by a personal grant of the Dutch Research Council (NWO; VENI: 09150161810095). A full list of acknowledgements for all the contributing studies can be found in the Supplementary Material table S1.

## Authors Contribution

EbvDA, DB, MJTR and PES conceived and wrote the manuscript. DB performed the analyses. EBvDA and MJTR verified and supervised the analyses. All authors discussed the results and contributed to the final manuscript.

